# Validation of a 9-item Perceived Suicide Awareness Scale (PSAS-9) for adolescents

**DOI:** 10.1101/2023.07.13.23292610

**Authors:** Stéphanie Baggio, Neslie Nsingi, Katia Iglesias, Marlène Sapin

## Abstract

**Introduction:** Robust empirical data on suicide awareness are needed, to better plan and evaluate suicide prevention interventions. However, there is a lack of validated measures of suicide awareness. This is especially true for perceived suicide awareness, which focuses on perceived knowledge about suicide, willingness, and confidence to talk about suicide and get help. This study aimed to validate a measure of perceived suicide awareness.

**Methods:** We re-used data from a suicide prevention trial conducted in Swiss secondary schools (n=366). Baseline and one-month follow-up data were used to validate the scale. The main measure was an initial 14-item perceived suicide awareness scale (PSAS). Perceived knowledge of help-seeking resources, suicide-related knowledge, and support networks were used to assess convergent validity.

**Results:** A nine-item version, the PSAS-9, showed satisfactory psychometric properties, including high internal consistency (α=.78), acceptable test-retest (r=.68), and a one-factor structure explaining 95% of the variance. The convergent validity was acceptable (.19≤r≤.40). The PSAS-9 was not correlated with suicide-related knowledge (r=.02).

**Conclusion:** This study was an important step towards validating a perceived suicide awareness scale, distinct from suicide-related knowledge, to be used in future studies focused on suicide prevention, and, more generally, studies interested in measuring suicide awareness.

## Introduction

Suicide is a leading cause of death among adolescents, and suicide prevention is a major public health need worldwide. Suicide prevention interventions encompass a wide range of potential interventions, including universal, selective, and indicated strategies (Goldsmith et al., 2002). Briefly, universal interventions target the entire population, whereas selective and indicated interventions focus on at-risk populations (those more likely to experience suicidal thoughts and behaviors) and high-risk individuals (those who have already experienced suicidal thoughts and behaviors), respectively. Universal interventions, the first step of prevention, are the core focus of this study. Universal interventions are programs designed to reach the greatest number of youth (Schwartz et al., 2022). Universal suicide prevention interventions often focus on suicide awareness, intending to improve knowledge (risk factors and warning signs) and attitudes (myths and preconceived ideas) about suicide, as well as reactions and help-seeking behaviors in case of suicidal behavior (Cusimano & Sameem, 2011).

Although suicide awareness is an essential outcome of universal suicide prevention, valid and reliable measures of this construct are lacking. Some validated scales measuring knowledge and attitudes are available, such as the Literacy of Suicide Scale (LOSS, Batterham et al., 2013). These scales include for example items such as “teenagers who talk about suicide do not kill themselves” or “If someone really wants to kill him/herself, there is not much I can do about it”. However, previous research often relied on non-validated scales and focus solely on knowledge and attitudes about suicide. Indeed, a recent systematic review of randomized controlled trials of universal suicide prevention programs in youth (Schwartz et al., 2022) only identified one study including suicide awareness as an outcome (Aseltine et al., 2007). It was based on a non-validated 48-item questionnaire assessing knowledge and attitudes about suicide (Shaffer et al., 1991; Spirito et al., 1988). In another recent systematic review of controlled trials (Brann et al., 2021), none of the studies that included suicide awareness as an outcome used a validated questionnaire.

Few measures are available for other dimensions of suicide awareness, such as improving reactions and help-seeking behaviors in case of suicidal behavior. Recent studies have used on a 14-item questionnaire (Baggio et al., 2022; Kinchin et al., 2020), developed by Bailey et al. (2017). This questionnaire tests perceived knowledge, confidence, and willingness to speak of suicide and get help and provides insights on “perceived suicide awareness”. It includes questions such as “I have the knowledge to recognize warning signs for suicide in others” (perceived knowledge), “I would be willing to seek help for suicidal thoughts” (willingness), or “I would feel confident enough to talk about suicide with others” (confidence). However, this questionnaire has also not been validated.

As suicide awareness is a critical outcome of universal suicide prevention interventions and to plan interventions at the population-based level, there is an urgent need for a validated questionnaire. Based on baseline data collected in a non-randomized universal suicide prevention trial (Baggio et al., 2022), this study aimed to validate a Perceived Suicide Awareness Scale (PSAS) that could be used in future studies focusing on suicide prevention and, more generally, in studies interested in measuring suicide awareness. As questionnaires are already available for the knowledge and attitudes’ part, it focuses on perceived knowledge, confidence, and willingness to speak of suicide and get help.

## Materials & methods

### Design and setting

This study was a secondary analysis of a non-randomized, cluster-controlled trial designed to test the effect of a brief suicide prevention program through workshops organized in secondary schools by the association *Stop Suicide* (Baggio et al., 2019; Baggio et al., 2022). The study took place in the French-speaking part of Switzerland, in several classes of two secondary schools located in Geneva and Neuchâtel, between December 2019 and November 2020. The trial included a baseline and a one-month follow-up. In this study, we used the baseline data of the intervention and control groups. We also used the follow-up data of the control group for the test-retest reliability. The study protocol was submitted to the cantonal ethics committee (no. 2019-00295) and was considered as falling outside of the scope of the Swiss legislation. Participants provided written informed consent.

### Participants

Inclusion criteria were 1) age 14 or older and 2) ability to communicate in French. The only exclusion criterion was having already participated in the *Stop Suicide* workshop in the previous year. Of 418 eligible adolescents, 373 agreed to participate (response rate = 90%). A total of 7 participants were excluded due to missing values on the perceived suicide awareness scale. The final sample consisted of 366 participants. The follow-up of the control group was used to assess the test-retest reliability. Of 100 control participants at baseline, 91 completed the follow-up (retention rate = 91%).

### Procedures

The association *Stop Suicide* (https://stopsuicide.ch) conducted a workshop on primary suicide prevention. It took place after the baseline assessment in the intervention group and after the one-month assessment in the control group. The data used in this study were collected prior to the intervention. At the baseline assessment, participants received information about the study and provided written consent. They then completed the baseline paper-and-pencil questionnaire (∼20 minutes). The same questionnaire, except socio-demographics, was used at follow-up.

### Measures

#### Perceived Suicide Awareness Scale (PSAS)

The 14 questions from a previous suicide prevention study were used to assess perceived suicide awareness (Bailey et al., 2017; Kinchin et al., 2020). The items deal with how people perceived their own knowledge and what are their attitudes towards suicide. It differs from objective knowledge of suicide, for which validated measures already exist. The initial PSAS includes five questions on perceived knowledge about suicide and help-seeking resources, three questions on willingness to talk about suicide and to get help, five questions on confidence to talk about suicide and get help, and one question on intention to get help. Items were rated on a five-point scale ranging from 0 = strongly disagree to 4 = strongly agree.

The scale was translated into French and then translated back into English. Discrepancies were discussed and resolved. The resulting scale was tested with the target population. The English questions are listed in Table 1.

**Table 1.** Descriptive statistics of the baseline assessment (n=366)

| Variables | Mean (sd) /<br>Percentage (n) |
| --- | --- |
| Age<sup>1</sup> | 15.3 (1.2) |
| Gender<sup>2</sup> |  |
| Boys | 44.0 (161) |
| Girls | 56.0 (205) |
| Parental level of education<sup>2</sup> |  |
| Primary or secondary | 54.1 (198) |
| Tertiary | 43.4 (159) |
| NA | 2.5 (9) |
| Perceived suicide awareness scale<sup>1</sup> |  |
| 1. I have the knowledge to recognize warning signs/invitations for suicide in myself. | 2.8 (1.0) |
| 2. I have the knowledge to recognize warning signs/invitations for suicide in others. | 2.0 (1.0) |
| 3. I have the knowledge to talk about suicide with others. | 2.4 (1.1) |
| 4. I have the knowledge to ask someone directly if they are thinking about suicide. | 1.8 (1.1) |
| 5. I have the knowledge to seek help for suicidal feeling. | 2.8 (1.0) |
| 6. I would feel willing to talk about suicide with others. | 2.6 (1.2) |
| 7. I would feel willing to ask someone directly if they are thinking about suicide. | 2.1 (1.1) |
| 8. I would be willing to seek help for suicidal thoughts. | 2.7 (1.1) |
| 9. I would feel confident when it comes to recognizing warning signs for suicide in myself. | 2.4 (1.1) |
| 10. I would feel confident when it comes to recognizing warning signs for suicide in others. | 2.2 (1.1) |
| 11. I would feel confident enough to talk about suicide with others. | 2.6 (1.1) |
| 12. I would feel confident enough to ask someone directly if they are thinking about suicide. | 1.2 (1.1) |
| 13. I would be confident enough to seek help for suicidal feelings. | 2.6 (1.2) |
| 14. How likely you would seek help for a problem like suicidal feelings? | 2.8 (1.1) |
| Knowledge of local help-seeking resources sum score (0-24)<sup>1,3</sup> | 15.3 (4.9) |
| LOSS (0-7)<sup>1</sup> | 5.4 (1.1) |
| Support networks<sup>2</sup> |  |
| Number of relatives | 3.1 (2.1) |
| Number of professionals | 0.7 (0.8) |

sd: standard deviation. ; LOSS: Literacy Of Suicide Scale.
<sup>1</sup> Means and standard deviations are given.
<sup>2</sup> Percentages and n are given.
<sup>3</sup> 5 missing values.

#### Perceived knowledge of help-seeking resources

We self-developed a scale to assess the perceived knowledge of local help-seeking resources. Six items were developed and scored on a five-point scale (ranging from 0 = strongly disagree to 4 = strongly agree). Items included, for example, “I know one phone number I can call to ask for help”. Items focused on potential local resources provided in the intervention (e.g., phone numbers and addresses, professionals at school and outside the school). A sum score was calculated, ranging from 0 to 24 (Cronbach α=.72).

#### Suicide-related knowledge

The French version of the Literacy of Suicide Scale (LOSS) was used to assess suicide knowledge and attitudes (Batterham et al., 2013). We used seven items from the original twelve-item scale, as other items were not covered by the intervention. A total score of correct responses from 0 to 7 was computed.

#### Support networks

Family and peer support networks moderate the relationship between psychological distress and suicide risk in adolescents (Thomas & Brausch, 2022) and may therefore be useful to assess convergent validity. We asked participants if they felt comfortable talking about their problems with 1) relatives (e.g., family members, friends), 2) classmates, and 3) professionals (e.g., general practitioner, specialist from a prevention league). Participants provided the first name and the detailed relationship to the person (e.g., mother, father, sibling, friend, neighbor, school nurse, teacher, psychologist). We calculated the number of support providers separately for relatives (categories 1) and 2) above) and for professionals (category 3) above).

#### Socio-demographics

Sociodemographic factors included gender, age, and parental education level (primary or secondary versus tertiary).

### Statistical analyses

We first ran descriptive statistics for the PSAS and all other variables using means (standard deviations) and percentages (n). We then divided the dataset into training and test sets, with a 70% vs. 30% split.

#### Analyses of the training set

We checked for item redundancy using the Goldbricker function. The Goldbricker function compares each pair of items and identifies measurement overlap by comparing correlation patterns with other variables in the dataset (topological overlap) (Payton, 2017). Redundant items (i.e., topological overlap greater than 75%) were removed. We tried to balance the number of items in each sub-dimension of PSAS (perceived knowledge, willingness, and confidence). We also reported the Pearson correlation matrix. We then tested the internal consistency using Cronbach’s alpha with a bootstrapped confidence interval. Finally, we performed an exploratory factor analysis (EFA) to identify the best factor structure of the PSAS.

#### Analyses of the test set

We performed a confirmatory factor analysis (CFA) using the factor structure retained from the EFA and the items selected from the training set. The Root Mean Squared Error of Approximation (RMSEA) and the Standardized Root Mean Squared Error (SRMR) were used to evaluate the model fit. Then, we tested the convergent validity using correlations between the PSAS and other related constructs (perceived knowledge of help-seeking resources, LOSS, and support networks). We expected these measures to be moderately related to the PSAS, as they investigated related, yet distinct, constructs. We used Pearson (perceived knowledge of help-seeking resources) and Spearman (LOSS and support networks) correlations. Finally, we calculated a Pearson correlation between the PSAS at baseline and follow-up in the control group for the test-retest reliability.

Participants were nested in classes. However, the intraclass correlation for the PSAS was low (0.03). Therefore, the clustering was omitted in the analyses. Because the PSAS items were normally distributed, they were considered continuous variables in all analyses. Statistical analyses were performed with R version 4.3.1 (packages lavaan version 0.6-15 and networktools 1.5.0).

## Results

Descriptive statistics are shown in Table 1. The mean age of participants was 15.3 ± 1.2 years, 56.0% were girls, and 54.1% had a primary or secondary parental level of education. Means of the PSAS items ranged from 1.2 (“I would feel confident enough to ask someone directly if they were thinking about suicide”) to 2.8 (“I have the knowledge to recognize warning signs of suicide in myself”, “I have the knowledge to seek help for suicidal feeling”, and “Indicate how likely would you seek help for a problem like suicidal feelings”). The means score of perceived knowledge of local help-seeking resources and LOSS were 15.3 and 5.4, respectively. Participants had, on average, 3.1 relatives and 0.7 professionals with whom they could speak of their problems.

### Analyses of the training set

The training set included n=260 (71% of the total sample). We first checked for item redundancy using the Goldbricker function. Less than 25% of the correlations were significantly different (i.e., topological overlap greater than 75%) for the pairs highlighted in bold in the correlation matrix shown in Table 2. We removed the redundant items while balancing the different sub-dimensions of the PSAS (perceived knowledge, willingness, confidence, and help-seeking behavior). Therefore, we removed the items 4 and 5 (knowledge), 12 and 13 (confidence), and 14 (help-seeking behavior). There was no item redundancy in the resulting 9-item PSAS (PSAS-9). The internal consistency of the PSAS-9 was α=.78 (95% confidence interval: .72; .82).

**Table 2.** Correlation matrix of the 14 items of the PSAS, training set (n=260)

|  | 1. | 2. | 3. | 4. | 5. | 6. | 7. | 8. | 9. | 10. | 11. | 12. | 13. |
| --- | --- | --- | --- | --- | --- | --- | --- | --- | --- | --- | --- | --- | --- |
| 1. | 1 |  |  |  |  |  |  |  |  |  |  |  |  |
| 2. | .177 | 1 |  |  |  |  |  |  |  |  |  |  |  |
| 3. | .175 | .354 | 1 |  |  |  |  |  |  |  |  |  |  |
| 4. | .141 | .386 | .326 | 1 |  |  |  |  |  |  |  |  |  |
| 5. | .239 | .182 | .193 | .244 | 1 |  |  |  |  |  |  |  |  |
| 6. | .157 | .193 | .409 | .327 | .342 | 1 |  |  |  |  |  |  |  |
| 7. | .165 | .403 | .282 | **.651** | .216 | .339 | 1 |  |  |  |  |  |  |
| 8. | .218 | .177 | .176 | .178 | **.536** | .368 | .187 | 1 |  |  |  |  |  |
| 9. | .259 | .202 | .199 | .217 | .242 | .251 | .142 | .299 | 1 |  |  |  |  |
| 10. | .055 | .343 | .239 | .322 | .200 | .307 | .364 | .241 | .414 | 1 |  |  |  |
| 11. | .167 | .263 | .472 | .316 | .176 | .604 | .342 | .251 | .371 | .409 | 1 |  |  |
| 12. | .156 | .349 | .340 | **.618** | .175 | .389 | **.765** | .211 | .146 | .370 | .427 | 1 |  |
| 13. | .179 | .124 | .174 | .128 | .536 | .399 | .150 | **.729** | .330 | .239 | .300 | .162 | 1 |
| 14. | .192 | .118 | .150 | .154 | .543 | .402 | .129 | **.742** | .351 | .229 | .285 | .131 | **.825** |

PSAS: perceived suicide awareness scale.
Items' labels are reported in Table 1. Items 1-5 are related to knowledge, 6-8 to willingness, 9-13 to confidence, and 14 to help-seeking behavior.
Correlations of redundant pairs of items are highlighted in bold.

The scree plot of the EFA is shown in Figure 1. It indicated that a one-factor solution was the best model. The first factor explained 95.5% of the total variance. Factor loadings for the PSAS-9 are shown in Table 3.

**Table 3.** Factor loadings of the PSAS-9 for the one-factor solution of the EFA, training set (n=260)

| Items of the SAS-9 | Loadings |
| --- | --- |
| 1. I have the knowledge to recognize warning signs/invitations for suicide in myself. | 0.27 |
| 2. I have the knowledge to recognize warning signs/invitations for suicide in others. | 0.44 |
| 3. I have the knowledge to talk about suicide with others. | 0.27 |
| 6. I would feel willing to talk about suicide with others. | 0.69 |
| 7. I would feel willing to ask someone directly if they are thinking about suicide. | 0.50 |
| 8. I would be willing to seek help for suicidal thoughts. | 0.42 |
| 9. I would feel confident when it comes to recognizing warning signs for suicide in myself. | 0.47 |
| 10. I would feel confident when it comes to recognizing warning signs for suicide in others. | 0.55 |
| 11. I would feel confident enough to talk about suicide with others. | 0.77 |

PSAS-9: Nine-item perceived suicide awareness scale; EFA: exploratory factor analysis.

**Figure 1.**
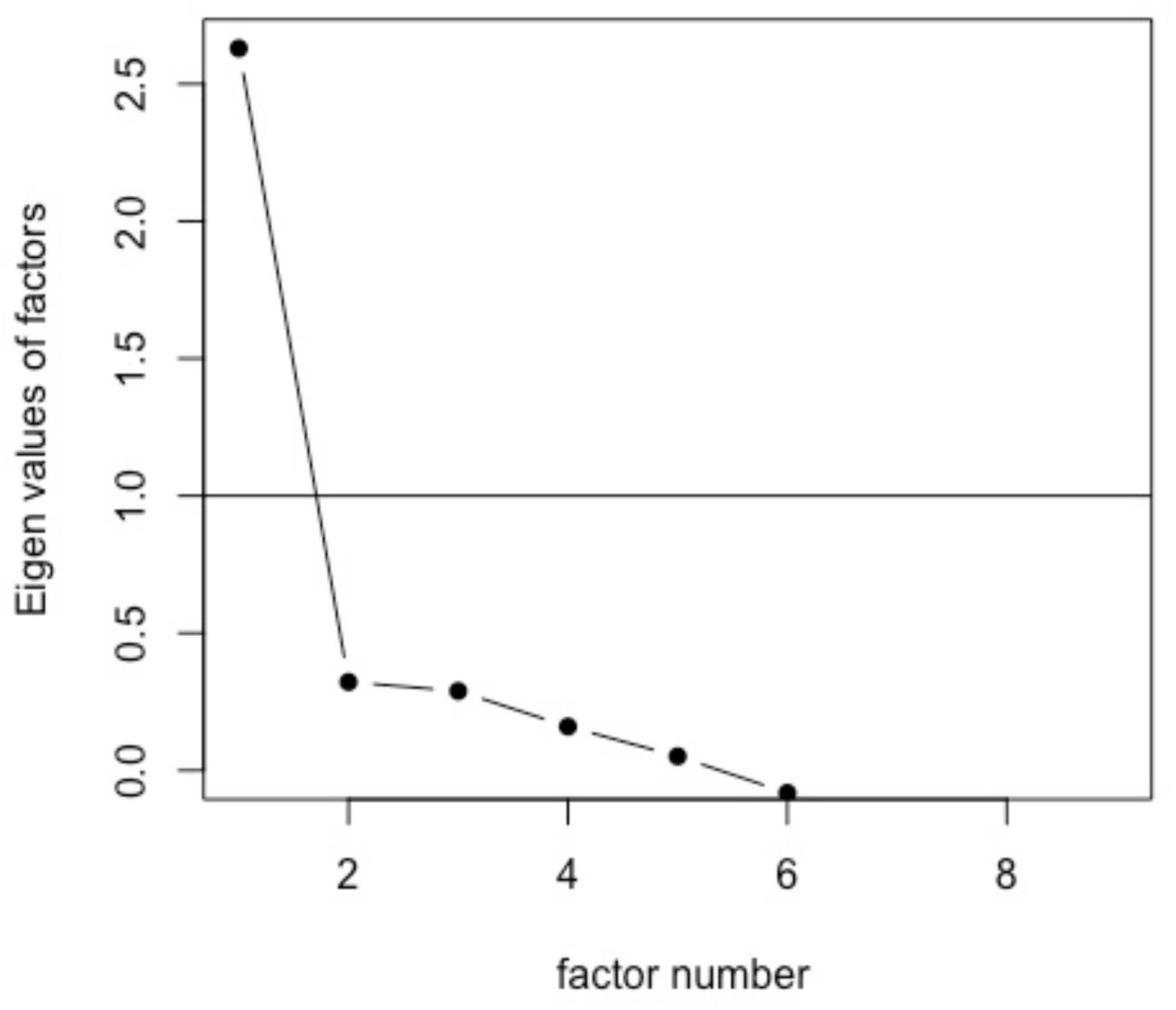
Scree plot of the exploratory factor analysis of the PSAS-9, training set (n=260) PSAS-9: Nine-item perceived suicide awareness scale.

### Analyses of the test set

The test set included n=106 (29% of the total sample). The internal consistency of the PSAS-9 was α=.78 (95% confidence interval: .70; .84). The one-factor CFA displayed acceptable psychometric properties for the SRMR (SRMR=0.069). The RMSEA was higher than expected (RMSEA=0.11). The standardized factor loadings are shown in Table 4. The PSAS-9 ranges from 0 to 36. The mean score of the PSAS-9 was 22.5 ± 5.6.

**Table 4.** Factor loadings of the PSAS-9 for the one-factor solution of the CFA, test set (n=106)

| Items of the SAS-9 | Loadings |
| --- | --- |
| 1. I have the knowledge to recognize warning signs/invitations for suicide in myself. | 1.00 |
| 2. I have the knowledge to recognize warning signs/invitations for suicide in others. | 0.83 |
| 3. I have the knowledge to talk about suicide with others. | 0.75 |
| 6. I would feel willing to talk about suicide with others. | 0.73 |
| 7. I would feel willing to ask someone directly if they are thinking about suicide. | 0.94 |
| 8. I would be willing to seek help for suicidal thoughts. | 1.03 |
| 9. I would feel confident when it comes to recognizing warning signs for suicide in myself. | 1.01 |
| 10. I would feel confident when it comes to recognizing warning signs for suicide in others. | 0.82 |
| 11. I would feel confident enough to talk about suicide with others. | 0.45 |

PSAS-9: Nine-item perceived suicide awareness scale; CFA: confirmatory factor analysis.

For convergent validity, the PSAS-9 had moderate positive correlations with perceived knowledge of help-seeking resources (r=.40, p<.001) and the number of people in the relatives’ support network (r=.32, p<.001). It had small positive correlations with the number of people in the professional support network (r=.19, p=.067). The PSAS-9 was not significantly correlated with the LOSS (r=.02, p=.764). The test-retest correlation of the PSAS-9 in the control group (n=91) was r=.68. (p<.001).

## Discussion

The purpose of this study was to validate a perceived suicide awareness scale, as there is currently a lack of validated scales to assess suicide awareness reliably. Perceived suicide awareness is defined as the perceived knowledge and attitudes (confidence, willingness to talk) toward suicide and help-seeking behaviors. It is defined as a different construct from objective knowledge and attitudes about suicide, for which validated measures already exist.

Building on a scale used in previous studies (Baggio et al., 2022; Bailey et al., 2017; Kinchin et al., 2020), we propose a 9-item Perceived Suicide Awareness Scale (PSAS-9). The PSAS-9 showed satisfactory psychometric properties, including high internal consistency, high reliability, acceptable test-retest, and a one-factor structure that can be easily used to derive a sum score ranging from 0 (low perceived suicide awareness) to 36 (high perceived suicide awareness), explaining 95% of the variance. The RMSEA was nevertheless higher than expected, suggesting that further research on the scale’s psychometric properties may be needed.

As no scale measuring the same construct exists, other constructs were used to evaluate the convergent validity. These constructs did not measure the same construct (i.e., perceived suicide awareness), but were related constructs. Therefore, small to moderate correlations were expected (correlations ≥ .5 would be expected when measuring the same construct). The convergent validity of the PSAS-9 was acceptable, with small to moderate correlations. The PSAS-9 had moderate positive correlations with the self-developed perceived knowledge of local help-seeking resources and the number of people in the relatives’ support network. The PSAS-9 had small positive correlations with the number of people in the professional support network. However, the number of professionals to whom participants could speak of their problems was reduced (on average, 0.7 per participant).

There was no link between the PSAS-9 and knowledge and attitudes about suicide, assessed with the LOSS. The correlation was .02. This suggests that, as expected, knowledge and attitudes about suicide and perceived knowledge, confidence, and willingness to talk and get help are different dimensions of suicide awareness. Of note, the level of knowledge and attitudes about suicide was high in the sample (mean=5.4 on a 7-point scale). Knowledge about suicide may therefore not be a sensible measure to test the benefit of interventions.

This study had some limitations. The first one was that the study was not originally designed to test the psychometric properties of the PSAS-9, so it did not include alternative measures of suicide awareness that could be used to assess the construct validity. Related constructs were used as proxies to evaluate the convergent validity, but the study missed a similar construct to properly assess convergent validity. A second limitation was the relatively modest sample size, resulting in a reduced power for tests of correlations. It led to marginally significant correlations for the convergent validity. A third limitation was that only secondary school adolescents were included in the analyses, so further research with other samples of youth in various research contexts is needed to robustly validate the PSAS-9 and confirm its factor structure. Finally, the LOSS was assessed using a subsample of original items, as the topics of the other items was not covered by the intervention. Future studies are needed to replicate the absence of correlation between perceived knowledge, confidence, and willingness to talk about suicide and get help and objective knowledge and attitudes about suicide. We suggest that both measures should be included as outcomes to test the benefits of suicide prevention interventions, as they do not overlap.

## Conclusion

This study was an important step towards validating a perceived suicide awareness scale, distinct from suicide-related knowledge, to be used in future studies focused on suicide prevention in various populations, and, more generally, studies interested in measuring suicide awareness.

## Data Availability

Anonymized data are available on reasonable request to the corresponding author.

## Notes

**Funding statement:** The work was supported by Promotion Santé Suisse.

**Conflict of interest disclosure:** The authors report no conflict of interest.

### Competing Interest Statement

The authors have declared no competing interest.

### Funding Statement

The work was supported by Promotion Sante Suisse.

### Author Declarations

The study protocol was submitted to the cantonal ethic committee (Geneva, no. 2019-00295). It was considered as falling outside the scope of the Swiss legislation (Federal Act on Research involving Human Beings).

## References

Aseltine, R. H., James, A., Schilling, E. A., & Glanovsky, J. (2007). Evaluating the SOS suicide prevention program: a replication and extension. BMC Public Health, 7(1), 161. https://doi.org/10.1186/1471-2458-7-161

Baggio, S., Kanani, A., Nsingi, N., Sapin, M., & Thélin, R. (2019). Evaluation of a suicide prevention program in Switzerland: protocol of a cluster non-randomized controlled trial. International Journal of Environmental Research and Public Health, 16(11), 2049. https://doi.org/10.3390/ijerph16112049

Baggio, S., Nsingi, N., Kanani, A., Bourqui, L., Graglia, M., & Thélin, R. (2022). Evaluation of a brief universal suicide prevention programme in young people: a cluster-controlled trial in Swiss schools. Swiss Medical Weekly, 152(2930), w30207–w30207. https://doi.org/10.4414/SMW.2022.w30207

Bailey, E., Spittal, M. J., Pirkis, J., Gould, M., & Robinson, J. (2017). Universal suicide prevention in young people: An evaluation of the safeTALK program in Australian high schools. Crisis: The Journal of Crisis Intervention and Suicide Prevention, 38(5), 300–308. https://doi.org/10.1027/0227-5910/a000465

Batterham, P. J., Calear, A. L., & Christensen, H. (2013). Correlates of suicide stigma and suicide literacy in the community. Suicide & Life-Threatening Behavior, 43(4), 406–417. https://doi.org/10.1111/sltb.12026

Brann, K. L., Baker, D., Smith-Millman, M. K., Watt, S. J., & DiOrio, C. (2021). A meta-analysis of suicide prevention programs for school-aged youth. Children and Youth Services Review, 121, 105826. https://doi.org/10.1016/j.childyouth.2020.105826

Cusimano, M. D., & Sameem, M. (2011). The effectiveness of middle and high school-based suicide prevention programmes for adolescents: a systematic review. Injury Prevention, 17(1), 43–49. https://doi.org/10.1136/ip.2009.025502

Goldsmith, S. K., Pellmar, T. C., Kleinman, A. M., & Bunney, W. E. (2002). Programs for suicide prevention. In Reducing Suicide: A National Imperative. National Academies Press (US). https://www.ncbi.nlm.nih.gov/books/NBK220931/

Kinchin, I., Russell, A. M. T., Petrie, D., Mifsud, A., Manning, L., & Doran, C. M. (2020). Program evaluation and decision analytic modelling of universal suicide prevention training (safeTALK) in secondary schools. Applied Health Economics and Health Policy, 18(2), 311–324. https://doi.org/10.1007/s40258-019-00505-3

Payton, J. (2017). Networktools: Assorted tools for identifying important nodes in networks. R package version 1.0.0. https://CRAN.R-project.org/package=networktools.

Schwartz, C., Yung, D., Barican, J., Gray-Grant, D., & Waddell, C. (2022). Suicide prevention: Reaching the greatest number of young people. Children’s Mental Health Research Quarterly, 16(4), 1–16. https://summit.sfu.ca/item/35225

Shaffer, D., Garland, A., Vieland, V., Underwood, M., & Busner, C. (1991). The impact of curriculum-based suicide prevention programs for teenagers. Journal of the American Academy of Child and Adolescent Psychiatry, 30(4), 588–596. https://doi.org/10.1097/00004583-199107000-00010

Spirito, A., Overholser, J., Ashworth, S., Morgan, J., & Benedict-Drew, C. (1988). Evaluation of a suicide awareness curriculum for high school students. Journal of the American Academy of Child & Adolescent Psychiatry, 27(6), 705–711. https://doi.org/10.1097/00004583-198811000-00007

Thomas, A. L., & Brausch, A. M. (2022). Family and peer support moderates the relationship between distress tolerance and suicide risk in black college students. Journal of American College Health, 70(4), 1138–1145. https://doi.org/10.1080/07448481.2020.1786096

